# Effects of nasogastric tube on oral microbiome among long-term care patients

**DOI:** 10.1101/2022.09.02.22279554

**Authors:** Ding-Han Wang, Fa-Tzu Tsai, Hsi-Feng Tu, Cheng-Chieh Yang, Ming-Lun Hsu, Lin-Jack Huang, Chiu-Tzu Lin, Wun-Eng Hsu, Yu-Cheng Lin

## Abstract

Dysbiosis of oral microbiome causes chronic diseases including dental caries and periodontitis, which frequently affects elderly, frail patients receiving long-term care. Severely disabled patients may require nutritional supply via nasogastric (NG) tube, which impacts patients’ oral condition and possibly microbial composition. However, little is known about the effect of NG tube on oral microbes and its potential ramification. Here, by using 16S rRNA next-generation sequencing, we characterized the tongue microbiome of 27 patients fed with NG tubes and 26 others fed orally. The microbial compositions of NG-tube and oral-feeding patients were substantially different, with more Gram-negative aerobes enriched in the presence of NG tube. Specifically, NG-tube patients presented more opportunistic pathogens like *Corynebacterium* and *Pseudomonas* associated with pneumonia, and lower levels of commensal *Streptococcus* and *Veillonella*. Together, we present a systematic, high-throughput profiling of oral microbiome with regards to NG tube indwelling, providing empirical evidence for better clinical practice.

**Importance:** Long-term use of NG tubes on elderly patients often leads to poor oral hygiene and chronic infectious diseases, e.g. periodontitis and tooth decay. More importantly, because patients fed with NG tubes usually have swallowing dysfunctions, they are more likely to suffer from aspiration pneumonia, a life-threatening lung infection caused by inhalation of oral bacteria. Together, clinical implications of chronic NG-tube indwelling are significantly related to oral microbes. Understanding the effects of NG tubes on oral microbiome would generally inform how clinical care should be given, particularly antimicrobial therapy.

## Introduction

The human oral cavity hosts hundreds of microbial species which play a crucial role in both oral and general health (1–3). Imbalanced oral microbiome, i.e. dysbiosis, is the primary causal factor of various oral and systemic diseases including dental caries, periodontitis (2), and infectious endocarditis (4). In addition, oral dysbiosis was shown to be associated with oral cancer, colorectal cancer, pancreatic cancer, rheumatoid arthritis, Alzheimer’s disease, and diabetes, yet definitive evidence for causal relationships is still lacking (2, 5). In the past, emergence of specific bacterial species was often viewed as the singular cause of chronic oral diseases; however, recent advancements in high-throughput sequencing technology have led researchers to realize that dysbiosis is a polymicrobial condition resulted from the complex interplay between the host and microbial community (6, 7). Oral dysbiosis often results in chronic illness which are particularly common in elderly, frail populations (8).

Elderly and disabled patients are more often to receive long-term care, which is a challenging situation for both caregivers and patients. One of the main factors affecting long-term care patients’ life quality is how nutrition is fed through the enteral route. Besides the normal oral intake, patients with cognitive impairment or swallowing difficulty require tube-feeding methods such as percutaneous endoscopic gastrostomy tube and nasogastric (NG) tube (9), the latter being the most common. During NG tube-feeding, patients’ normal oral function (e.g. chewing and swallowing) are significantly compromised. A dysfunctional oral cavity may lead to poor oral hygiene care (10) and negatively impact the oral microbial community, further complicating patients’ health condition. Although NG tube-feeding could be beneficial for the patients in the short term to deal with malnutrition (11), patients with prolonged NG-tube indwelling often suffered from dysphagia (12), malnutrition (13), and aspiration pneumonia (14).

Aspiration pneumonia, an acute condition caused by inhalation of oropharyngeal pathogens, poses major mortality risks to patients with NG tube as these patients often have impaired swallowing and cough reflux (15, 16). Aspiration pneumonia is most commonly treated with empiric antimicrobial therapy (17, 18), which in many cases could be ineffective due to the lack of information on the causative bacteria. Moreover, aspiration pneumonia is often caused by anaerobes which are hard to culture, further complicating the identification of the causative pathogen (16). Thus, systematic, culture-independent evaluation of pneumonia-associated oral microbes among long-term care patients is greatly needed for more precise antibiotics treatment.

Previous studies (19, 20) have addressed the effects of NG tube-feeding on oral microflora with older techniques, e.g. bacterial culture, restriction fragment length polymorphism, and pyrosequencing. In this work, we hypothesized that long-term indwelling of NG tubes alters the composition of oral microbiome, in a consistent and predictable manner. First, we sampled from the tongue as indicated by a previous study (21) that the tongue dorsum is the dominant source of microbes to cause aspiration pneumonia. Next, we used Illumina next-generation sequencing (NGS) targeting 16S rRNA amplicons to thoroughly detect the composition of tongue microbiome among NG-tube and oral-feeding patients. For data analysis, we re-designed our computational pipeline from the ground up, in order to achieve high resolution and accuracy in both taxonomic and diversity analysis. Specifically, we made improvements in (i) using amplicon sequence variant (ASV) (22) rather than operational taxonomic units (OTU), (ii) log transformation with pseudocount (23) for hierarchical clustering and beta diversity analysis, and (iii) simultaneous inclusion of two target sequences: V3 and V4 regions of the 16S rRNA (24). Together, these technical improvements enabled us to demonstrate that the oral microbiome of NG-tube and oral-feeding patients have significantly different microbial compositions. We also identified specific pathogens enriched in NG tube-feeding patients. Our work provides a global picture of the oral microbiome among NG-tube and oral-feeding patients and may better inform clinical care practices.

## Materials and Methods

### Patient recruitment and study design

This study involved 53 participants receiving long-term care from the Yilan County, Taiwan, and National Yang Ming Chao Tung University (NYCU) Hospital. Patients were included by the following criteria: (i) receiving long-term care due to moderate to severe disability, (ii) stable physical condition, and (iii) scheduled for routine dental care. Patients undergoing major medical intervention and antibiotics treatment were excluded. The recruitment period was from March to October 2021. 26 and 27 patients were nutritionally fed with oral feeding and NG tubes, respectively. Most patients fed with NG-tube resided in the community, receiving home-care dental service.

The study design and patient recruitment procedure (Fig. 1) was approved by the Ethics Committee of National Yang Ming Chiao Tung University (ID 2020B002). At the beginning of each patient’s recruitment, the procedure of the experiments was clearly explained to the participants and their family members, upon which informed consent was obtained. Questionnaire on basic clinical and demographic information on age, sex, systemic diseases, NG tube indwelling, and history of pneumonia were filled by either the participants or their family members. Then, detailed oral examination was performed by the dentist, followed by the collection of tongue plaque samples with sterilized cotton swabs.

**FIG 1.**
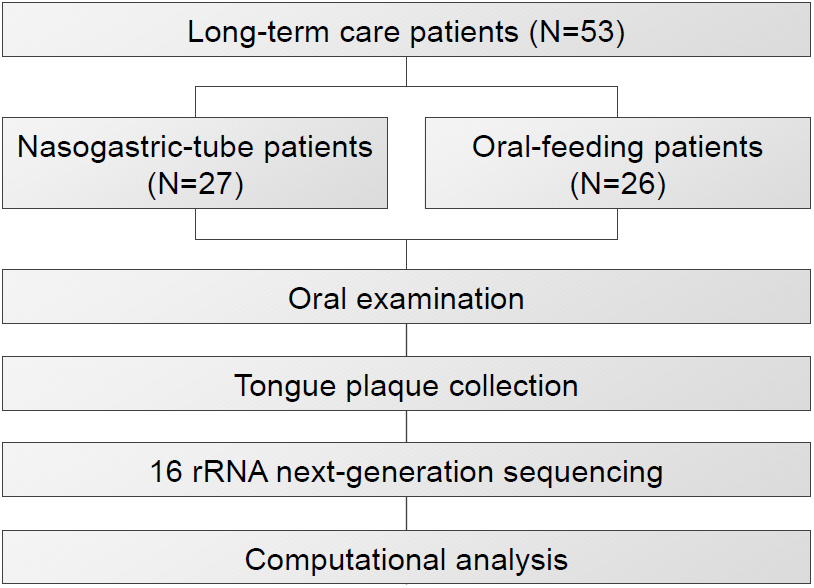
Flow chart of study design.

### Sample preparation and 16S rRNA amplicon sequencing

Tongue samples were collected using sterilized cotton swabs and stored in fresh, sterile TE buffer (10 mM Tris-HCl pH 8.0, 100 mM EDTA) at 4°C before being transported to the laboratory at AllBio Science Inc., Taichung, Taiwan for genomic DNA extraction within 24 hours. Total genome DNA was extracted from samples according to the manufacturer’s protocol (AllPure Bacteria Genomic DNA Kit). The quality of DNA concentration was controlled by Equalbit dsDNA HS Assay Kit. Samples were then stored at −20 °C for preservation before further target PCR amplification and sequencing steps. To avoid confounding batch effects, each batch consisted of 2 to 4 randomly assigned samples collected from either NG-tube or oral-feeding patients.

A segment covering hypervariable V3 to V4 regions of the prokaryotic 16S rRNA were used to generate amplicons, by using forward and reverse primers 341F (CCTACGGGNGGCWGCAG) and 805R (GACTACHVGGGTATCTAATCC), respectively (25). In the second-stage PCR, adapters (P5 and P7) and index sequences were added to either ends of amplified fragments. The DNA library was then purified with magnetic beads (AMPure XP), with concentration and fragment size measured by Qubit™ 3 Fluorometer (Thermo Fisher Scientific) and agarose gel electrophoresis, respectively. The library was adjusted to 10 nM before being sequenced with 150-bp paired-end sequencing on the Illumina MiSeq platform. 78,867 ± 38,367 and 77,290 ± 39,006 (avg ± SD) reads were sequenced for 27 NG-tube and 26 oral-feeding patients, respectively.

### Trimming and demultiplexing

The core of our computational pipeline was built on Qiime2 (26) version 2021.11 (https://docs.qiime2.org/2021.11/citation/), with custom numerical analysis implemented with Python (Fig. S1). To begin, paired-end reads were trimmed with cutadapt (27) wrapped by TrimGalore (https://github.com/FelixKrueger/TrimGalore) to remove primer sequences, low quality bases (minimum Phred quality 20), and reads that were too short (minimum read length 20 bp). Trimmed reads from multiple samples were then imported and demultiplexed as a compressed .qza file to enter the Qiime2 computational ecosystem.

### Denoising and taxonomic identification

Using Qiime2, Dada2 denoise algorithm (28) was used to detect ASVs, yielding a feature table (ASV × sample) and representative sequences for each ASV. Each representative sequence was taxonomically identified by using a pre-trained naïve Bayes classifier (https://data.qiime2.org/2021.8/common/silva-138-99-nb-classifier.qza) trained on the SILVA database (29). As each ASV was labeled with its predicted taxon, ASVs of the same taxon were pooled by summing their counts. The “taxonomic feature pooling” was performed at all taxonomic levels: phylum, class, order, family, genus, and species, yielding different levels of taxon tables (taxon × sample). Taken together, three data entities: (i) feature table (ASV × sample), (ii) taxon table (taxon × sample), and (iii) representative sequences were generated at this step for downstream analyses.

### Phylogenetic tree

Representative sequences for each ASV were compared using Mafft (30) multiple sequence alignment, generating a fasta alignment file, from which FastTree (31) was used to construct both rooted and unrooted phylogenetic trees in Newick (.nwk) format. The whole alignment/tree-building process was executed using the “qiime phylogeny align-to-tree-mafft-fasttree” command. The Python package ETE Toolkit (32) was used to visualize the phylogenetic tree (Fig. 3).

### Taxonomic composition

ASV feature counts were combined at all levels of taxonomy (phylum, class, order, family, genus, and species) by using Python packages “numpy” (33), “pandas”, and “matplotlib”. For each sample, we chose to show the quantity of the 20 most abundant taxa for visual clarity. All the remaining less-abundant taxa were pooled in the “Others” group (Fig. 4A).

### Hierarchical clustering

Clustering was performed on all levels of taxonomy (phylum, class, order, family, genus, and species). Prior to hierarchical clustering, the ASV feature or taxon counts underwent pseudocount (+1 for each count value) and then log10 transformation. Hierarchical clustering was performed by using the “clustermap” function of the “seaborn” package, with default parameters, i.e. “average” linkage method and “euclidean” distance metric (Fig. 4B).

### Alpha and beta diversities

From the feature table (ASV × sample), alpha diversities (Chao1 and Shannon index) (Fig. 2B) were computed using Qiime2. For beta diversity, Python packages including numpy (33), sklearn (34), and seaborn (35) were used. Briefly, log transformation with pseudocount was applied on the feature table prior to beta diversity analysis. Euclidean distance was chosen as the main distance metric for beta diversity. To visualize beta diversity on a 2D space (Fig. 5), principal component analysis (PCA) and t-stochastic neighbor embedding (t-SNE) were performed using the Python package “sklearn”, and subsequently plotted with “seaborn”. The “perplexity” parameter of t-SNE was manually fine-tuned to a value of 5. Analysis of similarity (ANOSIM) was performed using the Python package “scikit-bio”.

**FIG 2.**
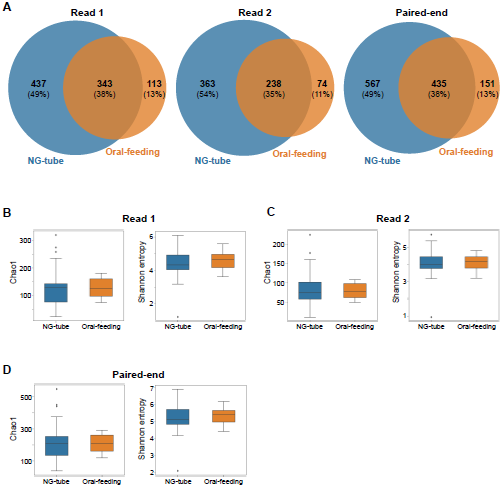
Consistent taxonomic composition and alpha diversity based on read 1 (V3 region) and read 2 (V4 region) sequences. (A) Venn diagrams of species detected from read 1, read 2, and pooled paired-end sequences. (B-D) Alpha diversity (Chao1 and Shannon entropy) computed from read 1 (B), read 2 (C), and pooled paired-end (D) sequences. The lower bound, middle line, and upper bound of a box mark the first quartile, median, and third quartile, respectively. Whiskers show the range from the smallest to the largest data points within mean ± 1.5 times of IQR (interquartile range). Points shown beyond the whiskers were considered outliers. No statistical significance of alpha diversity was detected between NG-tube and Oral-feeding patients.

### LEfSe analysis

LEfSe (36) version 1.1.01 was used. In brief, “lefse_run.py”, “lefse_plot_res.py”, “lefse_plot_cladogram.py” were executed in sequence to generate cladograms and LDA plots (Fig. 6). Like taxonomic composition analysis and hierarchical clustering, LEfSe was also performed on all taxonomic levels (phylum, class, order, family, genus, and species). We presented the LEfSe result on genus and species levels, as they provided the finest taxonomic resolution for differential abundance analysis.

### Spearman’s correlation

The Python package “scipy” was used to compute the correlation coefficient matrix and the corresponding *p* value matrix. *P* values were corrected with the Holm– Bonferroni method (37) by using the Python package “statsmodels”. Using adjusted *p* value ≤ 0.05 as the threshold, only significant correlation coefficient values were shown in Fig. 7 and Fig. S6.

### Software integration and interface

The complete data-analysis pipeline was fully integrated with Python. We provided an executable, user-friendly command-line interface (CLI). The source code is available at https://github.com/linyc74/qiime2_pipeline and containerized at https://hub.docker.com/repository/docker/linyc74/qiime2-pipeline.

### STORMS

The STORMS checklist (Mirzayi et al. 2021) can be downloaded at https://zenodo.org/api/files/c9fbbf13-4f00-4e38-9157-df538fa0fda8/STORMS%20Checklist.xlsx.

## Results

### Patient characteristics and oral health status

A total of 53 patients were recruited between March and October 2021 (Fig. 1). Table 1 summarizes all participants’ clinico-demographic background and oral health status. Notably, history of pneumonia was more frequently observed in the NG-tube (63%) than the oral-feeding (12%) group. In addition, patients fed with NG tubes presented higher caries index, residual root calculus, and more severe periodontitis. Overall, NG-tube patients presented worse general and oral health conditions compared to those fed orally.

**TABLE 1.** Patient characteristics and oral health status

|  |  | **NG-tube patients**<br>**(N=27)** |  | **Oral-feeding patients**<br>**(N=26)** |  |
| --- | --- | --- | --- | --- | --- |
| Age and Sex | Male/Female | 8/19 |  | 14/12 |  |
|  | Mean age (SD) | 73.92 (22.74) |  | 65.76 (16.77) |  |
| Disability level | Moderate | 1 | 4% | 7 | 27% |
|  | Severe | 13 | 48% | 14 | 54% |
|  | Profound | 13 | 48% | 5 | 19% |
| Diseases | Hypertension | 15 | 56% | 12 | 46% |
|  | Diabetes mellitus | 5 | 19% | 10 | 38% |
|  | Cardiovascular disease | 15 | 56% | 8 | 31% |
|  | Dementia | 8 | 30% | 5 | 19% |
|  | Liver disease | 5 | 19% | 1 | 4% |
|  | Kidney disease | 5 | 19% | 7 | 27% |
| Pneumonia history | Yes | 16 | 59% | 3 | 12% |
|  | No | 11 | 41% | 23 | 88% |
| Bedridden | Yes | 26 | 96% | 8 | 31% |
|  | No | 1 | 4% | 18 | 69% |
| Dental caries | Decay | 5 | 19% | 7 | 27% |
|  | Missing | 5 | 19% | 6 | 23% |
| Residual root | Yes | 12 | 44% | 11 | 42% |
|  | No | 15 | 56% | 15 | 58% |
| Periodontal status | Healthy | 0 | 0% | 4 | 15% |
|  | Gingivitis | 4 | 15% | 8 | 31% |
|  | Periodontitis | 23 | 85% | 14 | 54% |
| Calculus | Mild | 0 | 0% | 12 | 46% |
|  | Moderate | 1 | 4% | 4 | 15% |
|  | Severe | 26 | 96% | 10 | 38% |
| Dental Prosthetics | Partial | 0 | 0% | 2 | 8% |
|  | Full mouth | 0 | 0% | 3 | 12% |
|  | Crown / Bridge | 9 | 33% | 9 | 35% |

### Amplicon sequence variant as the primary feature

Although operational taxonomic unit (OTU) has been traditionally used as the fundamental feature for microbiome research, more recent developments in denoising algorithms such as dada2 (28) offers a better type of feature called amplicon sequence variant (ASV) (22). OTU suffers from low sequence specificity (usually at 97% similarity for species resolution), as sequencing errors could not be distinguished from true biological variation; in contrast, ASV can be resolved down to single-nucleotide level, exhibiting higher biological diversity without sacrificing data scalability (22). In this study, we used ASV as the fundamental feature of our analysis, and found detected ASV features (Fig. 3) greatly outnumbered the OTUs in our previous study (38). Using the ASV feature table as the core data structure, we developed a fully integrated computational pipeline that was executable with a single command line interface (Fig. S1).

**FIG 3.**
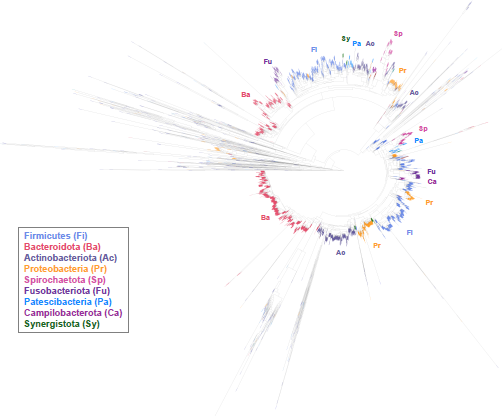
Phylogenetic tree constructed from 5,480 ASV features detected from pooled paired-end sequences covering V3 and V4 regions. Leaves were colored by predicted phyla, with the 9 most abundant phyla shown in the box. Around the perimeter of the circular tree, phylum clades were annotated with abbreviated names.

### Log transformation with pseudocount

Count-based analysis methods for high-throughput sequencing are often compounded by numerical overdispersion problems (39). Similarly, we observed overdispersed ASV counts, spanning 6 orders of magnitude (Fig. S2A), in which a few dominant ASVs overshadowed other less abundant, yet potentially important ASVs. Indeed, a similar overdispersion problem was also observed in bulk RNA-seq (40) and single-cell RNA-seq (41). To alleviate this count overdispersion problem, we applied log transformation with pseudocount (42) on ASV features, resulting in more evenly distributed abundance values that spanned only 2 orders of magnitude (Fig. S2B). The “fat tail” distribution by log-pseudocount made ASVs of all abundance levels more comparable, thus avoiding overdominance by a few extremely abundant ASVs. This evenness in log-pseudocount distribution was essential for subsequent hierarchical clustering (Fig. 4B) and beta diversity analysis (Fig. 5).

**FIG 4.**
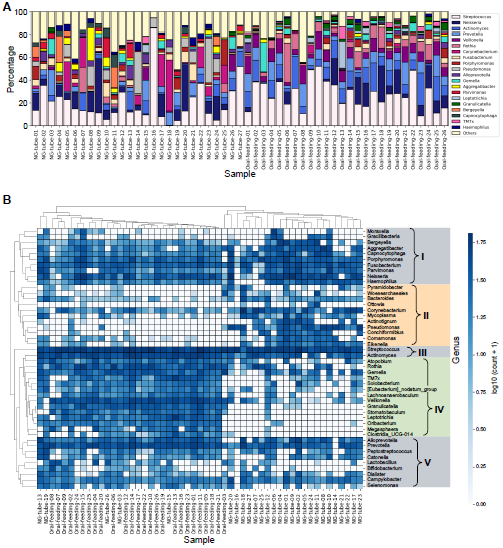
Genus composition and hierarchical clustering of the tongue microbiomes of NG-tube and oral-feeding patients. (A) Bar plot showing the relative abundance of annotated bacterial genera in each sample. Percentages were calculated by raw counts. The 20 most abundant genera across all samples were shown, with the remaining minor genera combined in the group “Others”. (B) Heatmap showing unsupervised hierarchical clustering of tongue samples and bacterial genera. Included were the 46 most abundant genera covering 95% of total reads. Five groups of genera were denoted from “I” to “V”. Color shades of the heatmap indicate log10 pseudocount values.

**FIG 5.**
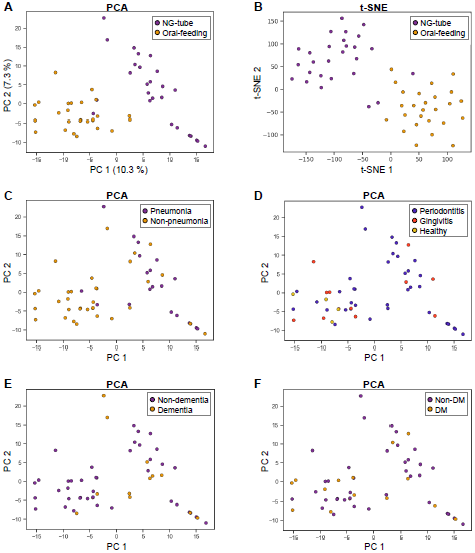
Beta-diversity analysis indicates distinct microbial composition between NG-tube and oral-feeding samples. Each point in the plot represents a tongue sample, with point-to-point distance indicating the Euclidean distance based on 5,480 ASV features after dimensionality reduction (PCA or t-SNE). (A, B) PCA and t-SNE plots colored by NG-tube vs. oral-feeding samples. Percentages in the parentheses indicate contributions of the two principal components, PC1 and PC2. (C-F) The same PCA plot in (A) colored by pneumonia history (C), periodontal status (D), dementia (E), and DM status (F).

### V3 and V4 regions yielded consistent taxonomic composition and alpha diversity

The choice of marker gene sequence (i.e. variable regions of 16S rRNA) is critical in resolving detailed microbial diversity and composition. To increase sequence diversity of the marker gene, we used 341F and 805R to amplify a 465 bp fragment that covers both V3 and V4 regions of 16s rRNA. 150 bp were sequenced with Illumina paired-end sequencing, with read 1 and read 2 covering V3 and V4 regions, respectively. As read 1 and read 2 do not overlap, paired-end reads cannot be merged as a single marker sequence, and should be treated as two separate markers.

We ran the computational pipeline separately on read 1 (V3 region) and read 2 (V4 region) sequences with identical configurations. Our result was consistent with a previous study on 16S sequence variability (24), that read 2 (V4 region) identified less species than read 1 (V3 region) (Fig. 2A). Nonetheless, the percentages unique to and shared by both feeding methods were consistent across read 1 and read 2 (Fig. 2A). Concordantly, detailed genus compositions of each sample were nearly identical between read 1 and read 2 (Fig. S3). As a consequence, alpha diversity (Chao1 and Shannon index) was consistent across read 1 and read 2, and showed no difference between NG-tube and oral-feeding groups (Fig. 2B-D). Together, our computational analysis performed robustly and consistently across the V3 and V4 regions of 16S rRNA.

Given the consistency between read 1 and read 2’s analyses, we pooled paired-end reads for subsequent computational analysis, to achieve maximum sequence diversity and taxonomic resolution. The Venn diagram based on pooled paired-end reads (Fig. 2A, right panel) have exactly the same percentages as those of read 1 (Fig. 2A, left panel), indicating the validity of the pooling approach. Pooling two target sequences would approximately double the amount of ASVs, achieving a total of 5,480 ASV features from 53 patient samples. The phylogenetic tree of all detected ASVs showed major phylum clades consistent with previously studied tongue microbiome (1, 20, 21, 43), e.g. Firmicutes, Bacteroidota, Actinobacteriota, and Proteobacteria (Fig. 4).

### Unsupervised hierarchical clustering showed distinct patterns of microbial composition between NG-tube and oral-feeding groups

Using pooled analysis on V3 and V4 sequences, we set off to examine detailed taxonomic compositions of tongue microbiome from NG-tube and oral-feeding patients. The 20 most abundant genera shown in Fig. 4A were commonly observed in previous studies, including *Streptococcus, Neisseria, Actinomyces, Veillonella, Corynebacterium, Fusobacterium*, and *Porphyromonas*. Notably, *Pseudomonas*, an opportunistic pathogen frequently associated with pneumonia (44), was among the most abundant genera as well (Fig. 4A).

We then examined whether NG tube and oral feeding has an effect on microbial composition. We converted the count of each genus to log-pseudocount, and applied unsupervised hierarchical clustering on the genera table (sample × genus). In the heatmap (Fig. 4B), distinct patterns were observed between NG-tube and oral-feeding samples, resulting in a nearly complete separation of two clades for the two feeding methods. We then assigned 5 major groups (I - V) of genera (Fig. 4B) based on their overall patterns across all samples. Group II and IV were enriched in NG-tube and oral-feeding samples, respectively. Generally, more Gram-negatives and a mixture of aerobes, facultatives, and anaerobes were found in group II genera (NG tube-associated), whereas group IV (oral feeding-associated) contained mostly Gram-positives anaerobes (Table S1).

Detailed inspection of the heatmap showed *Corynebacterium* and *Pseudomonas* were among the most overrepresented microbes in NG-tube patients (group II in Fig. 4B). Notably, *Corynebacterium* and *Pseudomonas* were both reported to cause aspiration pneumonia (44, 45), corroborating the fact that NG-tube patients had a higher incidence of pneumonia (Table 1). Conversely, group IV genera (enriched in oral-feeding patients) include *Veillonella, Leptotrichia*, and TM7x which were all common oral microflora (1, 43). Overall, distinct patterns of tongue microbial composition were clustered separately by NG-tube and oral-feeding lifestyles.

### Beta diversity showed distinct sample clusters correlated with feeding methods

The distinct patterns of NG-tube and oral-feeding samples in hierarchical clustering (Fig. 4B) led us to ask: To what extent do the tongue microbiomes differ from each other? Thus, we performed beta diversity analysis, i.e. sample-to-sample difference, using a total of 5,480 detected ASV features. We used Euclidean distance to measure the pairwise difference between samples. Next, we used PCA (principal component analysis) — a global, linear dimensionality reduction technique — to visualize the samples on a 2D plane. Strikingly, NG-tube and oral-feeding samples were almost completely separated (Fig. 5A), which is consistent with the finding in hierarchical clustering (Fig. 4B). In addition, t-SNE (t-stochastic neighbor embedding) (46) — a local, non-linear technique for high-dimensional data visualization — again showed that NG-tube and oral-feeding samples were separated (Fig. 5B). Finally, we performed analysis of similarity (ANOSIM) (47) to show that the difference between NG-tube and oral-feeding samples were statistically significant (*p* value = 0.0001) (Table 2). The positive value of test statistics (0.270579) indicates cohesion of intra-group samples relative to the inter-group samples (Table 2).

**TABLE 2.** Analysis of similarity (ANOSIM) on five categorical factors

| **Factor** | **Test statistics<sup>a</sup>** | ***p* value<sup>b</sup>** |
| --- | --- | --- |
| NG-tube / Oral-feeding | 0.270579 | 0.0001 |
| DM + / - | 0.008928 | 0.4498 |
| Dementia + / - | 0.023404 | 0.3945 |
| Pneumonia history + / - | 0.148019 | 0.0113 |
| Periodontitis / Gingivitis / Healthy | -0.03371 | 0.6118 |

<sup>a</sup>Positive values of test statistics indicate closer distance of intra-group samples compared to inter-group samples.
<sup>b</sup>*p* value was computed with 9,999 random permutations.

In addition to feeding methods, we wondered if the segregation pattern observed in the PCA plot could be attributed to other clinical factors. Sample points in the PCA plot were colored according to pneumonia history, periodontal status, dementia state, and diabetes mellitus (DM) (Fig. 5C-F). Among these factors, history of pneumonia was the only one that was separated by beta-diversity analysis, which could be due to the fact that NG-tube patients were correlated with a higher incidence of pneumonia (Table 1). Intriguingly, a previous study (48) showed that DM significantly altered subgingival microbiome in periodontitis, whereas in our study, DM had no effect on the tongue microbiome. Test results of ANOSIM (Table 2) were consistent with the observations in PCA plots, that pneumonia history was significantly associated with distinct microbial compositions (*p* value = 0.0113).

### Clinically relevant taxa were associated with NG-tube patients

Given that the tongue microbiome of NG-tube and oral-feeding patients have significantly different microbial composition (Fig. 4-5), we sought to identify specific taxa that were overrepresented in either NG-tube or oral-feeding patients. LEfSe (Linear discriminant analysis Effect Size) (36) was applied on all levels of taxon count tables (phylum, class, order, family, genus, species). Major clades of phyla were identified in the cladogram (Fig. 6A), in which Firmicutes and Actinobacteriota were associated with oral-feeding samples, while Proteobacteria, Spirochaetota, and Synergistota were associated with NG-tube samples.

**FIG 6.**
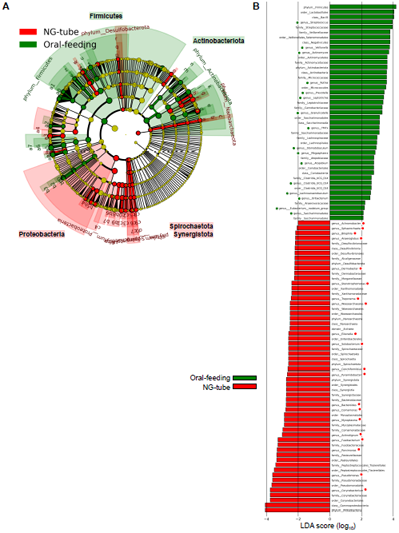
Genus-level LEfSe analysis revealed bacterial biomarkers enriched in NG-tube and oral-feeding patients. (A) Cladogram showing major bacterial phylum clades associated with NG-tube and oral-feeding patients. (B) LDA score of representative bacterial biomarkers from phylum to genus levels associated with NG-tube and oral-feeding patients. Red and green dots mark all genus-level biomarkers.

On the genus level, the two most associated genera in NG-tube patients were *Corynebacterium* and *Pseudomonas* (Fig. 6B), corroborating the findings in the heatmap (Fig. 4B). Importantly, both *Corynebacterium* and *Pseudomonas* have been reported as the causative pathogens of aspiration pneumonia (44, 45). Specific species of the two genera associated with NG-tube patients include *C. striatum, C. casei, P. stutzeri, P. aeruginosa*, and *P. otitidis* (Fig. S4). Surprisingly, the overrepresentation of *Corynebacterium* among NG-tube patients was also shown by Takeshita *et al. (20)*. In addition, genus *Parvimonas, Fusobacterium, Actinotignum*, and *Mycoplasma* were associated with NG-tube patients (Fig. 6B).

In oral-feeding patients, *Streptococcus* and *Veillonella* were highly enriched (Fig. 6B), which was again similar to the finding by Takeshita *et al. (20)*. The co-occurrence of the two genera was not coincidental, as studies (49, 50) have demonstrated a symbiotic relationship in which *Veillonella* metabolizes the lactate produced by *Streptococcus*, completing metabolic cross-feeding within a microbial community. Interestingly, interactions between *Streptococcus* and *Veillonella* have been mostly reported in dental plaque (51), whereas our study focused on the tongue dorsum. Additionally, genus *Actinomyces, Rothia, Prevotella, Leptotrichia, Granulicatella*, and TM7x were associated with oral-feeding patients (Fig. 6B). In conclusion, LEfSe identified pathogenic as well as commensal bacterial taxa associated with either NG-tube or oral-feeding patients.

### Co-occurrence analysis revealed antagonism between pathogenic and commensal species

We further asked whether the taxa associated with NG tube or oral feeding (Fig. 6) would co-occur with one another, given the fact that bacteria grow cooperatively or competitively within communities (7, 52). We particularly focused on species of the four genera: *Corynebacterium, Pseudomonas, Streptococcus*, and *Veillonella*, as they were either enriched or depleted in the presence of NG tubes (Fig. 6B). Spearman’s correlation coefficient was computed to measure pairwise co-occurrence of species. For visual clarity, we reordered the species in the correlation matrix such that more correlated ones were juxtaposed (Fig. 7). Two groups emerged, corresponding to commensals and pathogens associated with oral feeding and NG tube, respectively. The commensal group (Fig. 7; upper left) consisted of *Streptococcus* and *Veillonella* species, including *S. pseudopneumoniae, S. mitis, V. rogosae, S. sanguinis, V. atypica, S. salivarius, S. suis*, and *S. uberis*. In contrast, the pathogenic group (Fig. 7; bottom right) contained mostly *Corynebacterium* and *Pseudomonas*, including *P. otitidis, P. aeruginosa, C. striatum, C. simulans, C. casei*, and *P. stutzeri*. Interestingly, *Streptococcus infantis* was correlated with *P. otitidis* instead of the commensal Streptococci. Remarkably, negative correlation was observed between commensals (*S. sanguinis, V. atypica, S. salivarius, S. suis, S. uberis*) versus pathogens (*P. aeruginosa, C. striatum, C. simulans*) (Fig. 7; top right and bottom left). Taken together, co-occurrence was observed within respective commensal and pathogenic groups of bacteria, whereas an antagonistic relationship existed between pathogens and commensals.

**FIG 7.**
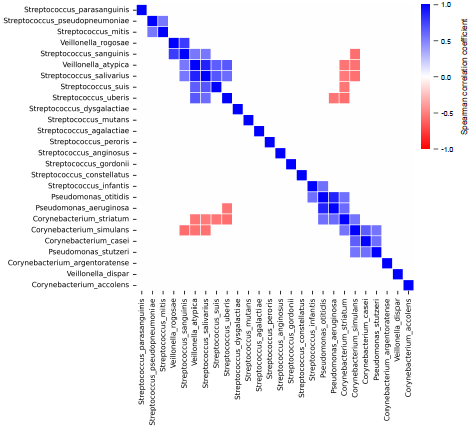
Co-occurrence of species of four genera: *Streptococcus, Veillonella, Corynebacterium*, and *Pseudomonas*, measured by Spearman’s correlation. Positive and negative correlation coefficients indicate co-occurrence and mutual exclusion, respectively. Only significant values were shown (Holm-adjusted p value ≤ 0.05).

## Discussion

In this study, we demonstrated that patients fed with NG tubes and those fed orally have substantially different tongue microbiomes. A conceptually similar study was conducted by Leibovitz *et al*. (19), in which oral microbes of elderly, frail patients were examined by using traditional culture-based techniques to detect pathogens. Although definitive, culture-based methods are inherently slow, low-throughput, and limited to culturable species. Our work presents a systematic, culture-free, and high-resolution profiling of oral microbiome regarding the effect of NG-tube indwelling. Such study could possibly inform the choice of antimicrobials before specific bacterial culture becomes available. Indeed, high-throughput sequencing has been used for fast, culture-free pathogen identification in various clinical scenarios (53–55).

A general notion exists that long-term indwelling of NG tube impairs swallowing function, thereby increasing the risk of aspiration pneumonia. Studies have shown that community-based elderly fed with NG tubes are generally at high risk of aspiration pneumonia (56). Moreover, pathogens of community-acquired pneumonia are generally different from those causing hospital-acquired pneumonia and thus require a different set of antimicrobials to treat (15, 57). Our survey of tongue microbes in patients with NG tubes showed mostly Gram-negative bacteria with a mixture of aerobes and anaerobes such as *Pseudomonas*, and also the Gram-positive *Corynebacterium* (Table S1). The spectrum of tongue microbes in NG-tube patients may provide predictive information for empiric antimicrobial therapy.

Challenges remain for microbiome study, one of them being the technical limitation to a small number of hypervariable regions of 16S rRNA. To resolve this problem, complete sequences of 16S rRNA need to be unambiguously defined, for which long-read sequencing technology such as PacBio SMRT has been applied (24). Nonetheless, even with the complete sequence of 16S rRNA, target genes are at best “proxy” for species identification, but not the comprehensive picture of physiologic and pathogenic functions. Virulence is essentially driven by microbial functions, not taxonomy. Supporting this notion, metabolic functions were shown to be well conserved across microbiomes from various body sites, despite radically different taxonomic compositions (43). With sequencing technology constantly becoming more powerful, shotgun metagenomic studies have emerged to functionally elucidate oral microbiome (58). Together, the use of long-read sequencing combined with shotgun functional metagenomics will shed new light on the complexity of the human microbiome.

## Conclusions

Our study in Taiwan presented strikingly similar findings to the study by Takeshita *et al*. (20) a decade ago in Japan, in which NG-tube patients harbored more opportunistic pathogens like *Corynebacterium* and less commensal *Streptococcus* and *Veillonella*. Cross-national evidence indicates that long-term NG-tube indwelling has predictable and deterministic effects on the oral microbiome, and thus indications for antimicrobial regimen.

## Data Availability

All data produced in the present study are available upon reasonable request to the authors

## Acknowledgement

The authors sincerely thank the support from the Department of Dentistry, National Yang Ming Chao Tung University Hospital, and all the participants in this study. This research is funded by the following grants: “110-2314-B-A49A-519”, “111-2314-B-A49-028-MY2”, “109-2314-B-010-012-MY2”, and “110-2314-B-A49A-551”, from the Ministry of Science and Technology, Taiwan. All authors declared no conflict of interest to disclose.

---

**FIG S1.**
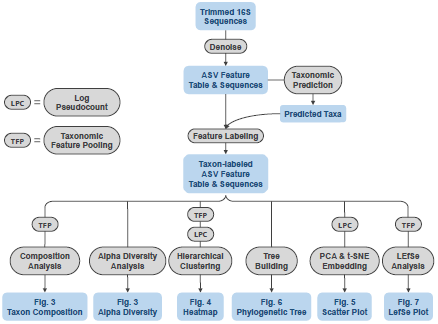
Computational pipeline diagram. Data structures and computational processes are colored in light blue and gray, respectively.

**FIG S2.**
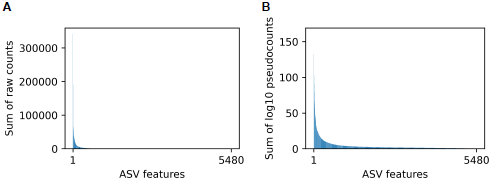
Ranked ASV feature counts showing sum of (A) raw counts and (B) log10 pseudocounts for each ASV for a total of 5,480 ASVs.

**FIG S3.**
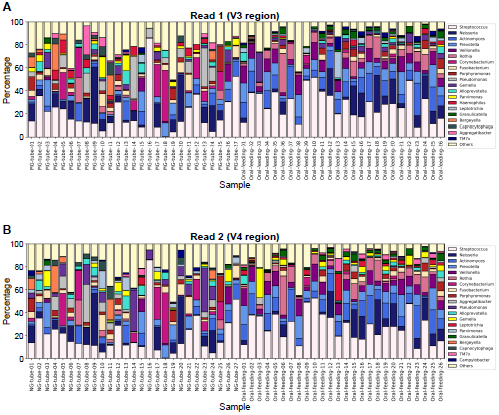
Genus composition detected with read 1 (V3 region) and read 2 (V4 region) sequences. The 20 most abundant genera across all samples were shown, with the remaining minor genera combined in the group “Others”.

**FIG S4.**
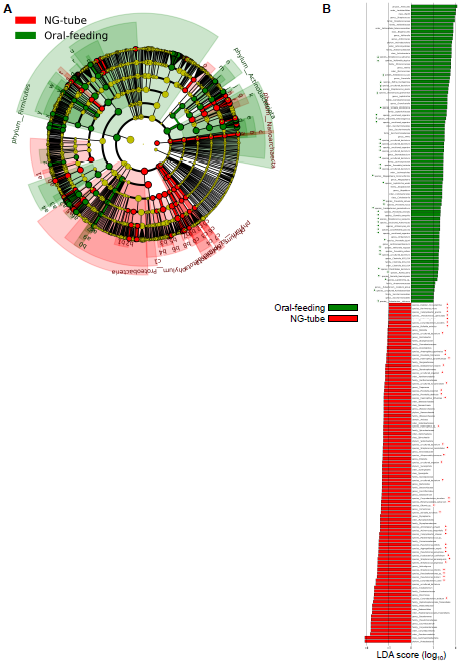
Species-level LEfSe analysis. (A) Cladogram and (B) LDA score showing bacterial taxa enriched in either NG-tube and oral-feeding samples. Red and green dots in (B) mark all species-level taxa.

**TABLE S1.** Gram stain and oxygen requirements of the genera shown in Fig. 4B

| Group | Genus | Gram stain | Oxygen requirements |
| --- | --- | --- | --- |
| I | *Moraxella* | - | aerobe |
| I | *Gracilibacteria* |  |  |
| I | *Bergeyella* | - | aerobe |
| I | *Aggregatibacter* | - | facultative |
| I | *Capnocytophaga* | - | facultative |
| I | *Porphyromonas* | - | anaerobe |
| I | *Fusobacterium* | - | anaerobe |
| I | *Parvimonas* | + | anaerobe |
| I | *Neisseria* | - | aerobe |
| I | *Haemophilus* | - | facultative |
| II | *Pyramidobacter* | - | anaerobe |
| II | *Woesearchaeales* |  |  |
| II | *Bacteroides* | - | anaerobe |
| II | *Ottowia* | - | aerobe |
| II | *Corynebacterium* | + | aerobe |
| II | *Mycoplasma* | - | facultative |
| II | *Actinotignum* | + | facultative |
| II | *Pseudomonas* | - | facultative |
| II | *Conchiformibius* | - |  |
| II | *Comamonas* | - | aerobe |
| II | *Eikenella* | - | facultative |
| III | *Streptococcus* | + | facultative |
| III | *Actinomyces* | + | facultative |
| IV | *Atopobium* | + | facultative |
| IV | *Rothia* | + | aerobe |
| IV | *Gemella* | + | aerobe |
| IV | *TM7x* | + | anaerobe |
| IV | *Solobacterium* | + | anaerobe |
| IV | *Eubacterium_nodatum_group* | + | anaerobe |
| IV | *Lachnoanaerobaculum* | + | anaerobe |
| IV | *Veillonella* | - | anaerobe |
| IV | *Granulicatella* | + |  |
| IV | *Stomatobaculum* | + | anaerobe |
| IV | *Leptotrichia* | - | anaerobe |
| IV | *Oribacterium* | + | anaerobe |
| IV | *Megasphaera* | - | anaerobe |
| IV | *Clostridia_UCG-014* | + | anaerobe |
| V | *Alloprevotella* | - | anaerobe |
| V | *Prevotella* | - | anaerobe |
| V | *Peptostreptococcus* | + | anaerobe |
| V | *Catonella* | - | anaerobe |
| V | *Lactobacillus* | + | facultative |
| V | *Bifidobacterium* | + | anaerobe |
| V | *Dialister* | - | anaerobe |
| V | *Campylobacter* | - | microaerophilic |
| V | *Selenomonas* | - | anaerobe |

